# Long-term Results of Combined Trabeculotomy-Trabeculectomy in Primary Congenital Glaucoma from a Tertiary Eye Care Center in Tunisia

**DOI:** 10.1101/2022.08.11.22278657

**Authors:** Ines Malek, Jihene Sayadi, Racem Choura, Manel Mekni, Haythem Rayhane, Moncef Khairallah, Leila Nacef

## Abstract

**Objective:** To report the long-term visual and surgical outcomes of combined trabeculotomy–trabeculectomy (CTT) as the initial glaucoma surgery in children with primary congenital glaucoma (PCG).

**Methods:** Prospective analysis of children who underwent primary CTT for PCG between January 2010 and December 2019. The main outcome measures were intraocular pressure (IOP) reduction, corneal clarity, success rate, complications, refractive errors and visual acuities (VA).

**Results:** A total of 98 eyes of 62 patients were enrolled. At last follow-up, mean IOP reduced from 22.68±3.99 mmHg to 9.75±3.88 mmHg (P<0.0001). Complete success rate was 91.6%, 88.4%, 84.7%, 71.6%, 59.7%, and 54.3%, respectively at first, second, fourth, sixth, eighth, and tenth year. Follow-up averaged 42.06±28.36 months. Preoperatively, 72 eyes (73.5%) had significant corneal haze, 84.7% among them achieved normal corneal transparency. Among the sight-threatening complications, endophthalmitis was encountered in one eye. Myopia was the most common refractive error (80.6%). Data on Snellen VA were available for 53.2% of the patients. 33.3% of patients achieved normal VA (VA ≥ 6/12), 21.2% had mild visual impairment (VI), 9.1% had moderate VI, 21.2% had severe VI, and 15.2% were blind. The failure rate was statistically correlated to the early disease onset (<3 months) and to preoperative corneal clouding (p=0.022 and p=0.037, respectively). Two critical time points were identified: within the first year and around the sixth year (28.6 and 25% of surgical failures, respectively).

**Conclusion:** Primary CTT seems to be an optimal procedure in a population with advanced disease at presentation, problematic follow-ups, and limited resources.

**Summary box:** *What is already known on this topic:*

- Primary congenital glaucoma (PCG) is the most common type of childhood glaucoma. Surgery is the main therapeutic option with the goal of permanently controlling IOP and preserving visual function. Several surgical procedures are available. Each approach has its potential benefits and risks. The optimum first-line surgery is debated.

*What this study adds?:*

- PCG in Tunisian children seems to be characterized by a high prevalence of inherited and advanced form of the disease.
- Challenges in the management of PCG in Tunisia include remoteness of care facilities and poor compliance to follow-up.
- Primary combined trabeculotomy-trabeculectomy allowed in the current study satisfactory long-term IOP control and reasonable visual outcome. The overall incidence of serious complications was low.

**How this study might affect research, practice or policy:**

- CTT may be the optimal primary surgical procedure for the management of PCG in Tunisia and all other developing countries. Future research with a larger sample size and preferably randomized clinical trials that focus exclusively on the severe presentation of the disease are required.

## Introduction

Primary congenital glaucoma (PCG) is a major cause of childhood blindness worldwide [1]. Although rare, it is a severe disease characterized by elevated IOP and subsequent optic neuropathy leading to irreversible visual defects [1-4]. Due to its huge impact, PCG requires early diagnosis, prompt treatment, and follow-up examinations throughout patients’ lives [1-4]. PCG is responsible for 0.01-0.04% of blindness worldwide [4]. This condition accounts for 0.7-4% of all new glaucoma cases in sub-Saharan regions of Africa [5]. However, it would occur up to 10 times more often in some ethnic groups where consanguineous marriage is more prevalent [5]. In India, it is responsible for 4.2% of severe childhood visual impairment [6]. In Tunisia (North Africa), 18.8% of childhood blindness is due to PCG [7].

Currently, the only therapeutic option of PCG is surgical with the goal of permanently controlling IOP and preserving visual function [1-4]. Goniotomy and trabeculotomy are considered initial procedures among the glaucoma specialists in the Western populations because of their high success rates and low incidence of complications [2-4,6,8-10]. However, in many developing countries children with PCG often present with familial and advanced form of the disease with severe degree of corneal clouding [4-7,11] in which goniotomy is technically unfeasible, and undertaking only ab externo trabeculotomy would result in poor IOP control [4,6,11]. Furthermore, challenges in the management of PCG in limited resources countries include remoteness of care facilities and poor compliance to follow-up [4-6,11].

Owing to these racial, social and economic particularities, the combined trabeculotomy-trabeculectomy (CTT) as a “first-line treatment” and hopefully “the single operative procedure” for PCG tends to become the preferred surgical option in several developing countries [4,6, 12-20]. In addition, this practice has proven to be safe and effective in numerous studies [4,6,12-21].

Therefore, we conducted this study to assess long-term surgical and visual outcomes as well as factors influencing the surgical success of CTT in children with PCG operated by a single surgeon in a North African pediatric population.

## Methods

### Study design

A prospective study was conducted at the Hedi Rais Institute of ophthalmology of Tunis, Tunisia, which is a tertiary referral center. Eligible patients were recruited between January 2010 and December 2019. The study adhered to the tenets of the Declaration of Helsinki. The use of CTT as the initial therapeutic modality was approved by the Ethics Committee of Aziza Othmana Hospital. A written informed consent was obtained from the parents after thoroughly explaining the benefits and risks of the surgical procedure.

The study included patients with PCG who underwent CTT as initial glaucoma surgery. We did not include children with secondary glaucoma, patients with a history of previous surgery for PCG and patients with coexisting ocular diseases such as congenital corneal dystrophy, congenital iris abnormality, congenital cataract or retinopathy of prematurity. All surgeries were performed by a single surgeon (IM).

According to the Childhood Glaucoma Research Network classification of PCG patients were subcategorized into three groups based on age of onset: neonatal or newborn (0– 1 month), infantile (>1–24 months), and late-onset glaucoma (>24 months) [22,23].

Patients with bilateral disease underwent delayed sequential bilateral glaucoma surgeries within one week.

### Preoperative examination

Detailed demographic data were elicited, including family history of PCG, parental consanguinity, geographic origin, perinatal and neonatal history, presenting complaints, delay in seeking glaucoma care and anti-glaucoma medications.

All patients underwent a complete preoperative ocular examination under anesthesia (EUA) to assess the disease and its baseline severity.

EUA was performed under sevoflurane anesthesia using spontaneous ventilation. IOP was measured in the early phase of anesthesia after induction using a hand-held Perkins applanation tonometer (Clement Clarke International Ltd., Essex, UK). IOP >16 mmHg under general anesthesia was considered as abnormal. [4,6,24]

Corneal clarity was evaluated under high magnification and HAAB striae were detected. Horizontal corneal diameter (HCD) was measured using Castroviejo calipers. Cycloplegic refraction (using 1% cyclopentolate) and gonioscopy (using Koeppe’s gonioscopic lens, Ocular Instruments, Inc., Bellevue, WA) were performed as permitted by the corneal clarity. Fundus examination to assess optic disc changes (measurement of vertical cup-to-disc (C/D) ratio) was done using indirect ophthalmoscopy. When fundoscopy was precluded, ultrasound B-scan was performed to detect excavation of the optic nerve head and to rule out any intraocular pathology.

### Surgical procedure

Under general anesthesia, a fornix-based conjunctivo-tenon flap was raised and a mild hemostasis was achieved using wet field cautery. Afterwards, a 4-mm partial thickness limbus based triangular scleral flap was dissected. A 2×2-mm trabeculectomy flap was marked along the bottom of the scleral flap. A 2 mm central radial incision was then performed across the scleral spur and slowly deepened until the external wall of Schlemm’s canal opened up, at which point there was a seeping of aqueous humour. Once Schlemm’s canal identified, trabeculotomy was performed. The internal arm of the trabeculotome was rotated into the anterior chamber, and the inner wall of Schlemm’s canal and the trabecular meshwork were dissected 100° to 120° in both directions. After the trabeculotomy was completed, trabeculectomy was performed by removing the deeper block marked previously. A peripheral iridectomy was performed. The triangular scleral flap was sutured with one suture at the apex using 10-0 nylon sutures. Finally, the tenon layer was sutured with separate 8-0 polyglactin stitches while the conjunctiva was closed in a continuous fashion using the same suture.

### Postoperative treatment and patient follow-up

A standardized postoperative care regimen was followed including dexamethasone eye drops 6 times a day for the first week, followed by a slow taper over the next 7 weeks; topical gentamycin was given 4 times a day for 2 weeks.

All patients were examined at the 1^st^, 5^th^ and the 15^th^ day to observe for immediate postoperative complications including hyphema, shallow anterior chamber, hypotony, iatrogenic cataract, retinal or choroidal detachment, and endophthalmitis. EUA in young infants was performed at the 1^st^, 3^rd^, 6^th^,9^th^ and 12^th^ month then each 6 months until the patient became old enough to be examined without anesthesia.

Postoperative EUA included measurement of IOP, bleb characteristics, HCD, evaluation of corneal transparency, lens clarity, C/D ratio and refractive error. If the patient’s IOP was found to be high, appropriate anti-glaucoma medications were prescribed and closer monitoring was scheduled.

Amblyopia therapy was immediately initiated. We emphasized the importance of wearing glasses and patching.

Best-corrected visual acuity (BCVA) was measured in cooperative children using various age-appropriate charts. We reported BCVA using Snellen charts only. Visual acuity was reported according to the International Classification of Diseases (11^th^ edition, 2018) criteria for vision loss, as used by WHO, which categorizes people according to vision in the better eye on presentation.

In the current study, we reported the visual acuity (VA) in the affected eye in unilateral cases and the VA in the better eye in bilateral cases. The categories of visual impairment (VI) were as follows: mild VI (VA of 6/18 or better but less than 6/12), moderate VI (VA of 6/60 or better but less than 6/18), severe VI (VA of 3/60 or better but less than 6/ 60), and blindness (VA of less than 3/60). Patients whose VA was below 3/60 were referred to the vision rehabilitation centers.

### Success criteria

In our study, we adopted the success criteria of Mandal and coauthors studies [6, 14, 16, 17]. Success was defined as:

- A final IOP ≤16 mmHg under anesthesia or a final IOP ≤ 21mmHg if the patient is old enough to be examined without anesthesia
- Without progression of the corneal diameter; and
- Without progression of the optic disc cupping.

Success was deemed “complete success”, when the above-mentioned end-points were achieved without need for antiglaucoma medication. Otherwise, it was deemed “qualified success”.

CTT was considered as failure if the above-mentioned objectives were not achieved despite medical therapy or if the patient developed hypotony-related maculopathy or sight-threatening complications, or required additional glaucoma surgery.

### Statistical analysis

IBM SPSS Statistics version 23 was used for the statistical analysis. In the descriptive statistics, we calculated means and standard deviations for quantitative variables, and absolute and relative frequencies for qualitative variables. In order to compare preoperative to postoperative data, we used a paired t-test for means and a Pearson’s Chi2 test for frequencies.

Kaplan-Meier survival curves have been used to assess the success rate after CTT surgery. The identification of risk factors linked independently to the failure rate was processed by multivariate analysis in logistic regression with the step-by-step method.

In all statistical tests, the significance level was set at 0.05.

## Results

### Demographic data

Our study group included 98 eyes of 62 patients with PCG who underwent CTT as the initial glaucoma surgery. Follow-up was prospective averaging 42.06 ± 28.36 months (range, 6-120 months; median, 36 months).

Parents’ consanguinity was elicited in 28 patients (45.2%). Eleven patients (17.7%) had a family history of congenital glaucoma. Of our patients, 36 (58.1%) came from the north-west of the country.

There were 28 male children (45.2%) and 34 females (54.8%), defining the sex-ratio at 0.82. Buphthalmos was the main presenting complaint in 23 patients (37.1%), followed by corneal clouding in 19 patients (30.6%), excessive tearing and sensitivity to light in 15 patients (24.2%), and red eyes in 5 patients (8.1%). Fourteen patients (22.6%) had newborn glaucoma, 36 (58.1%) had infantile glaucoma and 12 (19.3%) had late-onset glaucoma. The disease was bilateral in 36 patients (58.1%) and unilateral in 26 patients (41.9%). The mean age at surgery was 5.89 months (range, 1.5–33 months).

### Intraocular pressure

The mean preoperative IOP was 22.68 ± 3.99 mmHg (range, 18–32 mmHg). Of our patients, 45 patients (72.6%) were already on anti-glaucoma medications at the time of referral.

At the last follow-up, the mean IOP was 9.75 ± 3.88 mmHg (range, 4–26 mmHg). The mean IOP reduction was 12.93 ± 5.16 mmHg, the percentage reduction in IOP was 56.06% and was statistically significant (p<0.0001).

### Corneal parameters and C/D ratio

Preoperatively, only 17 of the 98 eyes (17.3%) had normal corneal transparency. Seventy-two eyes (73.5%) had varying degrees of corneal edema. Five eyes (5.1%) presented with hydrops and 4 eyes with corneal scarring (4.1%). HAAB’s striae were observed in 22 eyes (22.4%).

At the last follow-up visit, 78 eyes (79.6%) had normal corneal transparency, 3 eyes (3.1%) had corneal edema and 17 eyes (17.3%) had corneal scarring. Among the 72 eyes with preoperative corneal oedema, 61 (84.7%) recovered a corneal transparency. The corneal status improvement was statistically significant (p<0.0001).

At presentation, 37 eyes (37.8%) had an HCD above 14 mm. Mean HCD was 13.47 ± 0.92 mm (range, 11.0–15.0 mm). At the last follow-up, mean HCD was 13.54 ± 0.89 mm (range, 11.5–15.0 mm) (p=0.857).

Thirty-four out of 98 eyes (34.7%) had clear enough media for optic disc assessment preoperatively with a mean C/D ratio of 0.53 ± 0.24. The mean C/D ratio at last follow-up of the same 34 eyes was 0.54 ± 0.27 (p=0.912).

A summary of the demographic data and clinical parameters was given in Table 1.

**Table 1:** Demographic profile and clinical features of the study population.

| Characteristics | Results |
| --- | --- |
| **Gender** |  |
| Male | 28 (45.2%) |
| Female | 34 (54.8%) |
| **Consanguinity** | 28 (45.2%) |
| **Type of glaucoma** |  |
| Neonatal (0–1 month) | 14 (22.6%) |
| Infantile (>1–24 months) | 36 (58.1%) |
| Late-onset (>24 months) | 12 (19.3%) |
| **Horizontal corneal diameter at presentation (mm)** |  |
| ≤14 | 61 (62.2%) |
| > 14 | 37 (37.8%) |
| **Corneal clarity at presentation** |  |
| Clear | 17 (17.3%) |
| Oedema | 72 (73.4%) |
| Hydrops | 5 (5.1%) |
| Scarring | 4 (4.1%) |
| HAAB striae | 22 (22.4%) |
| **Mean time between symptom's onset and surgery (days)** | 4.12 (1 to 12) |
| **Age at surgery (months)** | 5.89 (1.5 to 33) |
| **Corneal clarity at last visit** |  |
| Clear | 78 (79.6%) |
| Oedema | 3 (3.1%) |
| Scarring | 17 (17.3%) |
| **IOP** |  |
| Preoperative IOP (mmHg) | 22.68 ± 3.99 (range, 18–32 mmHg) |
| Postoperative IOP (mmHg) | 9.75 ± 3.88 (range, 4–26 mmHg). |
| Reduction of IOP (%) | 56.06 % |
| **Mean Follow-up (months)** | 42.06 ± 28.36 (range, 6-120 months) |

### Complications

Peri-operative complications were encountered in 16 eyes (16.3%). **(Table 2)**

**Table 2:** Peri-operative complications and adverse events in study population.

| **Complications** | **n (%)** |
| --- | --- |
| Hyphema (required wash-out in one eye) | 11 (11.2%) |
| Shallow Anterior Chamber (restored spontaneously in all cases) | 3 (3.1%) |
| Iatrogenic Cataract | 1 (1%) |
| Endophthalmitis | 1 (1%) |
| **Total** | **16 (16.3%)** |

Hyphema was the most common early post-operative complication, observed in 11 eyes (11.2%). Chamber wash-out was required in one eye. A shallow anterior chamber was noted in 3 eyes (3.1%). It was restored spontaneously in all cases.

Iatrogenic cataract occurred in one eye among the first performed surgeries. It was related to the trabeculotomy learning curve. VA regained baseline after cataract surgery.

An endophthalmitis occurred in one eye. The causative factor was the corneal stay sutures used during the surgical procedure. It was successfully managed by intravenous (cefotaxime and fosfomycine), and intravitreous injections of antibiotics (vancomycine and ceftazidime).

There was no choroidal detachment, rhegmatogenous retinal detachment, bleb leakage or bleb-related infection.

### Refractive errors and visual acuity

Data regarding refractive errors were available for 72 eyes (73.5%). Fifty-eight eyes (80.6%) were myopic with mean SE of -6.6 ± 4.13 D (range, -1.25 to -19.25 D). Eight eyes (11.1%) were hyperopic with mean SE +2.2 ± 0.51 D (range, +1 to +4.25D) and 6 eyes (8.3%) were emmetropic.

Snellen acuity measurement were available for 33 patients (53.2%). Eleven patients (33.3%) achieved normal VA (VA ≥ 6/12), 7 (21.2%) had mild VI, 3 (9.1%) had moderate VI, 7 (21.2%) had severe VI, and 5 (15.2%) were blind as defined by WHO criteria of vision loss.

### Success rate

The success rates within the ten years follow-up are summarized in Table 3. Figure 1 represents the Kaplan-Meier survival curves for complete success and complete plus qualified success. Seventeen patients (27.4%) were lost to follow-up within the three first years. Two critical time points were identified: within the first year (28.6% of surgical failures) and around the sixth year (25% of surgical failures).

**Table 3:** Success rate of eyes with primary congenital glaucoma

| Follow-up duration | Success rate % (95% CI) |  |
| --- | --- | --- |
|  | Complete | Complete + Qualified |
| 6 months | 98 (96.6 – 99.4) | 99 (98 – 100) |
| 1 year | 91.6 (88.8 – 94.4) | 93.7 (91.2 – 96.2) |
| 2 years | 88.4 (85.1 – 91.7) | 92.6 (89.9 – 95.3) |
| 3 years | 87.3 (83.9 – 90.7) | 91.5 (88.6 – 94.4) |
| 4 years | 84.7 (80.9 – 88.5) | 89 (85.7 – 92.3) |
| 5 years | 83.3 (79.3 – 87.3) | 89 (85.7 – 92.3) |
| 6 years | 71.6 (66.3 – 76.9) | 82.5 (78.1 – 86.9) |
| 7 years | 65.1 (59.1 – 71.1) | 78.5 (73.5 – 83.5) |
| 8 years | 59.7 (53.1 – 66.3) | 76.1 (70.7 – 81.5) |
| 9 years | 54.3 (46.4 – 62.2) | 72.5 (66.3 – 78.7) |
| 10 years | 54.3 (46.4 – 62.2) | 72.5 (66.3 – 78.7) |

**Figure 1.**
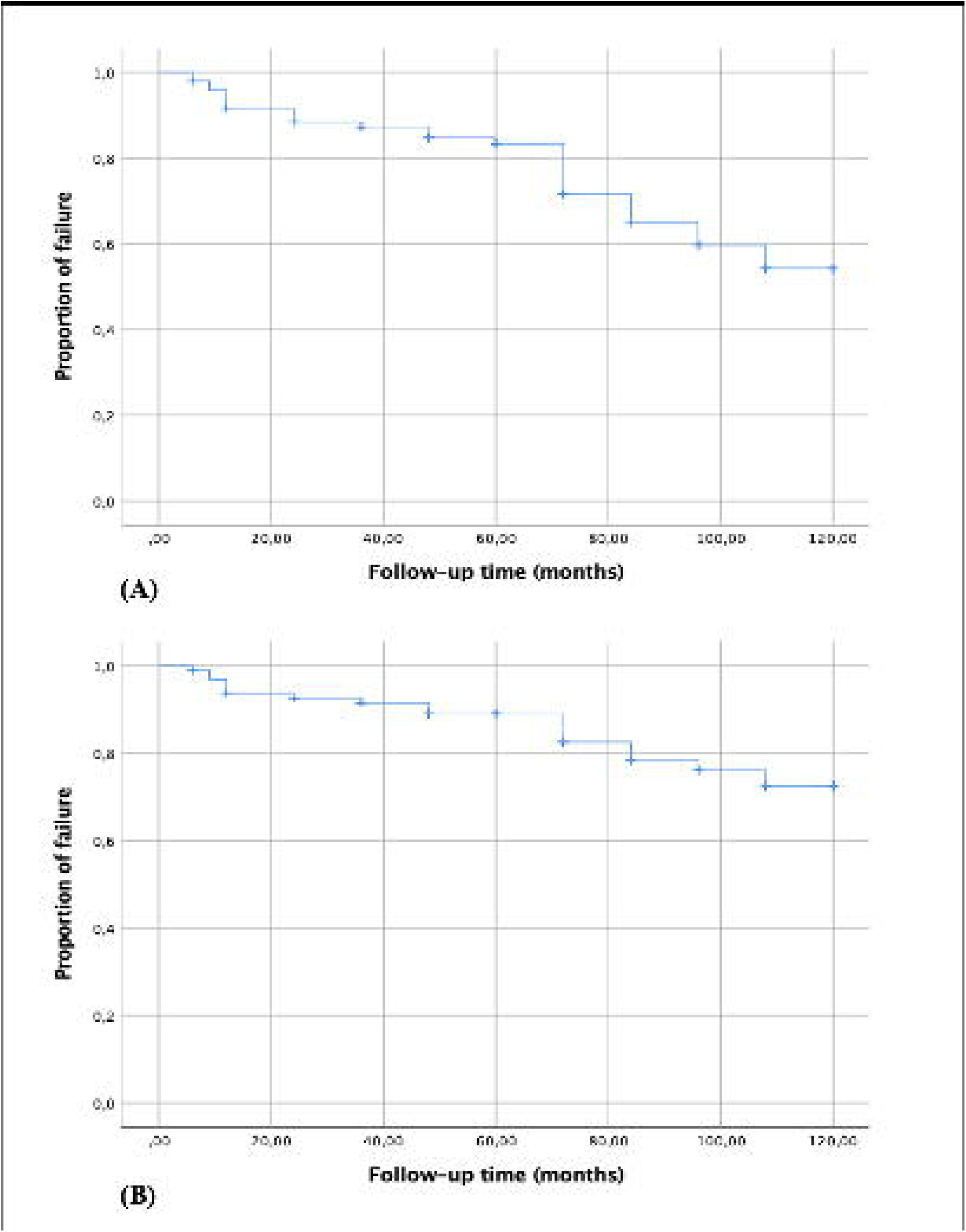
Kaplan–Meier survival curves showing the success probabilities of combined trabeculotomy-trabeculectomy (CTT) for our study population in 98 eyes: (A) complete success. (B) complete plus qualified success.

On multivariate analysis, failure rate was statistically correlated with the early disease onset (<3 months) (p=0.022) and preoperative corneal clouding (p=0.037). However, it was not associated with gender (p=0.642), preoperative IOP (p=0.498), preoperative HCD (p=0.511) or time between onset of symptoms and surgery (p=0.713).

## Discussion

Despite being the most prevalent glaucoma in infancy, there have been few African reports focusing on long-term outcomes of PCG surgery [15, 20,25-27]. According to our series, PCG in Tunisian children seems to be characterized by a high prevalence of familial and advanced form of the disease. In our cohort, inherited PCG accounted for 45% of cases. Most of our patients (58.1%) came from the north west side of Tunisia. This region is known by the high prevalence of consanguineous marriages and therefore a higher risk for autosomal recessive diseases, including PCG. Approximately the third of our children (37.75%) showed an HCD above 14 mm at presentation, HCD being an indicator of the disease severity [17,28].

Similar to many developing countries, there are several issues and challenges associated with PCG management in Tunisia: lack of disease awareness and fear of surgical treatment, difficulties to reach the health care facilities, presentation of the disease in an advanced state, inadequate follow-up, and the limited number of specialized surgeons in PCG due to the condition rarity [5,11].

In our country, simultaneous bilateral CTT should be considered especially in patients with problematic health care access. The second eye surgery should be performed as if it is being undertaken on a different child with strict aseptic conditions [8,16].

Several surgical procedures have been used to treat PCG. However, the optimum first line approach remains subject of considerable debate especially that final outcomes would highly depends on the initial glaucoma surgery results [6,14,17,29].

CTT emerged as a promising treatment modality which may overcome the limitations of conventional surgeries and yield superior results [12-15]. Combined procedures create a dual exit pathway for aqueous: through the Schlemm’s canal in trabeculotomy and through the trabeculectomy fistula [17]. It is suggested that trabeculectomy allows the bulk flow of aqueous during the early postoperative period, whereas the opened trabeculotomy cleft allows the long survival of CTT [18].

The most persuasive argument favoring primary CTT was the higher success rate with a single procedure [6,12-17]. In the current study, CTT resulted in significant IOP reduction in PCG, with a complete success rate of 91.6% at the end of first year follow-up, 87.3% at the third-year that declined to 54.3% at the end of the tenth year.

In a large and recent study by Mandal and coauthors including 1128 eyes of 653 patients who underwent bilateral CTT, Kaplan-Meier survival analysis revealed 1-, 5-, 10-, and 19-year complete success rates of 92.6%, 75.5%, 55.9%, and 21.6%, respectively [6]. Our success rates over ten years of follow-up appear to be very similar to these study results as well as those from previous studies using CTT as primary procedure [12,14,18, 19, 24,30]. Furthermore, our results of long-term success are yet better than those reported by Kessel and coauthors. in a recent series of 96 PCG children (144 eyes) with a follow-up of 35 years. Ab externo trabeculotomy was the first-line surgery in their study [10].

It is reasonable to expect that the higher success rate obtained by combining trabeculotomy to trabeculectomy would be associated to a higher complications rate. Nevertheless, studies revealed that main CTT complications are only due to overfiltration risk including hypotony, shallow anterior chamber, and their related long-term sequelae [6,14,16, 28]. Taking these complications into account, we did not use adjunctive mitomycin-C (MMC). Similarly, Mandal and coauthors did not use adjunctive MMC. When comparing their surgical results with those reported in studies using MMC, they concluded that the application of MMC in primary CTT would not improve the surgical success rates [6].

The major complication we have encountered was endophthalmitis. The limbal stay suture was the causative factor. Since this occurrence, we used superior rectus traction suture as an alternate method of globe traction.

Consistent with previous data, the earlier disease onset and preoperative corneal clouding were strongly associated with failure [6, 10, 31, 32]. Mandal and coauthors have shown other important risk factors including a preoperative IOP>35 mmHg and a history of previous glaucoma surgery. When both factors coexisted the risk of failure was 6 times higher [33].

One of the objectives of the present study was to clarify whether glaucoma progression was clustered around specific time points. Two critical time points were identified: within the first year and around the sixth year following surgery. Zagora and coauthors identified three critical time points: within the first year of life, around the 2-to-3-year and 5-to-6-year age groups [9]. Nonetheless, they performed regular angle surgeries as the primary procedure. Increased vigilance during critical time points is recommended.

Myopia was the commonest refractive error following glaucoma surgery in our series as well as in the series of Mandal and coauthors [6,14,17,33]. High incidence of myopia is attributed to the advanced nature of the disease and the young age at presentation. In addition to early and successful surgery, appropriate refractive correction combined to aggressive amblyopia therapy is mandatory to reach an optimum visual rehabilitation [1,2,5,11].

In developing countries, a high rate of children lost to follow-up is commonly reported (27.4% within the first three years in our series) [6,11]. Snellen acuity data were available in only 53.2% of our patients. Reasons of poor visual outcome in children with complete success include corneal scarring, HAAB striae, irregular astigmatism, anisometropic or high ametropic amblyopia, and optic nerve damage [4,6].

To summarize, in Tunisia, the majority of the children with PCG present with inherited and severe form of the disease. The overall incidence of serious complications was low in our series and the achieved IOP control and visual benefit were reasonable. Thus, CTT may be the optimal first-line surgical procedure for the management of PCG in Tunisia and all other developing countries.

There are several limitations to the current study. First, the study included a relatively small single-center series. Second, the lack of complete VA data, the precise assessment of VA being technically challenging in very young children. Third, a high rate of patients was lost to follow up (27.4% within three years from surgery). However, to the best of our knowledge, this series is the first single-center study to date assessing the long-term surgical as well as visual outcome of primary CTT in the Maghreb region.

In conclusion, our study demonstrated over a long-term, that CTT allows an excellent IOP control with no severe complications at reasonable cost. Thus, based on this study and review of literature, CTT seems to be safe, efficient and predictable enough to be preferable to a regular angle surgery in a population with advanced disease at presentation and problematic follow-up. However, further investigation including multicenter prospective studies with lower rate of patients lost to follow-up is needed to better assess the efficacy and safety of CTT in treatment of PCG.

## Data Availability

All data produced in the present study are available upon reasonable request to the authors

## Declaration of interest

The authors report no conflicts of interest. None of the authors has a financial or proprietary interest in any material or method mentioned. This work has not been supported by any public or private institution.

## Notes

### Competing Interest Statement

The authors have declared no competing interest.

### Funding Statement

This work has not been funded by any public or private institution.

### Author Declarations

The Ethics Committee of Aziza Othmana Hospital, Tunis, Tunisia approved this research.

